# Leadership and governance of mental healthcare and integration at the community level: a mixed methods study in Ghana

**DOI:** 10.1101/2023.11.09.23298301

**Authors:** Peter Badimak Yaro, Emmanuel Asampong, Philip Teg-Nefaah Tabong, Graham Thornicroft, Paulina Tindana

**Affiliations:** Department of Health Policy, Planning and Management, School of Public Health, Box LG 13, College of Health Sciences, University of Ghana, Legon, Accra; Department of Social and Behavioural sciences, School of Public Health, BOX LG 13, College of Health Sciences, University of Ghana, Legon, Accra; Health Service and Population Research Department, Institute of Psychiatry, King’s College London, London.

## Abstract

Leadership and governance are key components of health systems, nevertheless research into leadership and governance of mental healthcare at the community level is probably the least well researched and understood part of these systems. As part of assessing the integration of mental health in Primary Health Care in Ghana, the leadership and governance organisation and structures to ensure oversight and coordination were examined. A concurrent mixed-methods design involving both quantitative and qualitative research methods approach was adopted. The quantitative data were collected through a questionnaire, which was either self-administered or interviewer administered, on 1010 respondents with 830 completed (response rate 82%). Key informant interviews and focus group discussions were used to collect the qualitative data. Thematic content analysis with the use of NVivo 12 was applied for the qualitative field data and Stata SE16 was used for quantitative data. Data triangulation strategy was used to report the qualitative and quantitative results. The study showed that leadership and governance of mental health at the PHC level were lowly developed due to the modest level of awareness of the Mental Health Law, inadequate functioning of mental health units and coordination, low level of private sector participation in mental health care services, and low levels of provision of monitoring, supervision, and evaluation. This affected the integration of mental health at the PHC level, which was also gauged as low. The study concludes that despite the presence of legislation and policy aiming to achieve decentralised and integrated mental health services at the PHC level, mental health care is still a low-level priority within the health care system in Ghana and tends to operate within a silo. The study recommends that more practical and concerted leadership of mental health at the regional and district levels is required to drive decentralisation and integration at these levels.

## Introduction

Only a small proportion of people living with mental health conditions receive treatment despite the existence of effective public mental health interventions for the treatment of mental conditions, prevent associated impact and increase of mental health conditions, and promotion of mental health and wellbeing overall (1–3). The scale-up of mental health care and people-centred community-based mental health care service models are best achieved through integration into general healthcare at the Primary Health Care (PHC) level (4–6). Integration of mental healthcare into general healthcare at the Primary Health Care (PHC) level remains the core and singular global consensus to scaling-up mental healthcare and addressing the high treatment gap in resource limited settings, especially in Low– and Middle-Income Countries (LMICs) (Lund, Tomlinson and Patel, 2016; World Health Organization [WHO], 2016; Gwaikolo, Kohrt and Cooper, 2017).

Integration of mental health care within the general health care system in PHC is best appreciated within the context of the interaction of the components of the Health System Framework proposed by the WHO and further enhanced by the Ouagadougou Declaration on Primary Health Care and Health Systems in Africa (Member States of the African Region of WHO, 2008; WHO, 2007). The WHO Health System Framework is bringing together an interaction of the various building blocks that constitute a health system to be able to deliver health care services to meet the health needs of the population (12). The key building blocks of the health system include: (i) Leadership and Governance; (ii) health service delivery; (iii) health workforce/ health human resources; (iv) health information systems; (v) health products, vaccines and technologies; (vi) health financing; (vii) community ownership and participation; (viii) partnership for health development; and (ix) research for health (13).

In recent years, the growth in understanding and appreciation of the importance of mental health in population health and wellbeing has heightened the need to develop mental health systems to respond to the mental health care service needs (Gouda et al., 2019; WHO, 2019). Leadership and governance are important components of the health system framework in the development and integration of mental health policy and services at the community level. They are considered the hub around which the various components of the health system framework revolve (13). The WHO succinctly sets out that leadership and governance involves ensuring the existence of a strategic policy framework which combines “effective oversight, coalition-building, regulation and attention to system-design and accountability” (16). It takes the interactions of three categories of stakeholders to determine the health system and its leadership/governance (17–19). These include the state, the healthcare service providers, and the citizens, particularly service users. ‘State’ covers the government Ministries, Departments and Agencies (MDAs) at the central and regional and local levels. ‘Healthcare service providers’ refer to public and private for profit and not-for-profit clinical providers, para-medical and non-medical healthcare providers, unions, and medical/ health professional associations, as well as networks of care or of services. The third category of stakeholders in the health system leadership and governance is the ‘citizens’, particularly population representatives such as user/patient associations, and Non-Governmental Organisations (NGOs) and Civil Society Organisations (CSOs).

Essential to effective leadership and governance for integrated mental health system are legislation, policies, and strategies for ensuring a coordination and oversight (20–22). Ghana’s mental health law, mental health policy, and strategic plan affirm the global understanding of what a health system thinking is (23–25). Key provisions of the Mental Health Law (Act 846; 2012) and the overall purpose and interpretation of the legislation emphasises decentralisation of mental healthcare services down to the community level, including the integration of mental healthcare services into general healthcare services at the PHC level. The expected decentralisation and integration of mental health care into general care at the community level in Ghana, remains rather limited in practice and far less successful overall (26,27).

Mental health care service provision in Ghana remains largely a silo in the healthcare delivery arrangements of the country, especially at the community level, perpetuating the disconnection, isolation, and discrimination of mental health in general health care services across the country (28). The bulk of resources for mental health care (financial, human and logistical/ medicines) go to the three psychiatric hospitals of Ghana, located in the southern-coastal regions, rather than the regional, district and lower levels of healthcare delivery which would serve a bigger population than the three psychiatric hospitals do. This situation has contributed to the low resourcing and investments in mental health at the lower levels of the healthcare system, and a generally low priority accorded mental health.

A most likely problem of such situation with mental health in Ghana could be leadership and governance inadequacies. Since the passage of the Mental Health Act (Act 846), in 2012, there has not been a critical examination and clarification of the roles and responsibilities of the Mental Health Authority (MHA) and the Ghana Health Service (GHS) in the coordination/ management structure, organization, and delivery of mental health care services outside of the psychiatric hospitals, for that matter at the Primary Health Care level. The MHA draws heavily from the Mental Health Act (846) to operate while the GHS substantially relies on the Public Service Act, 1996 (525) to deliver its mandate of providing health care services as a public service body (29,30). These two legally backed roles have somewhat affected the way and manner community mental health services are organized, managed, and delivered. The Mental Health Act, 2012 (846) was promulgated to address the years of neglect of the sub-sectors affecting the organisation of services and overall oversight and leadership in the country (31). A decade following the promulgation of this Act in Ghana and nearly eight years of the coming into being of the MHA operating along with the GHS, the intended structure and organization of mental health services outside of the psychiatric hospitals have remained blurred. Improving and sustaining community mental health is integral to overall health system strengthening efforts.

There is, therefore, the interest and need to examine the key dynamics and indicators that could promote integration of mental healthcare at the PHC level of the Ghana’s healthcare system. The aim of this study was therefore to assess the leadership and governance structures put in place to ensure oversight, coordination, and general stewardship of mental healthcare at the PHC in Ghana.

## Materials and methods

### Study Design

This study adopted a concurrent triangulation mixed-methods design (32–36). It involved the use of both qualitative and quantitative approaches (37–39).

### Study area

The study was conducted in the Republic of Ghana. Ghana is divided into sixteen regions (Figure 1) and three ecological zones (Northern, Middle, and Southern).

**Figure 1:**
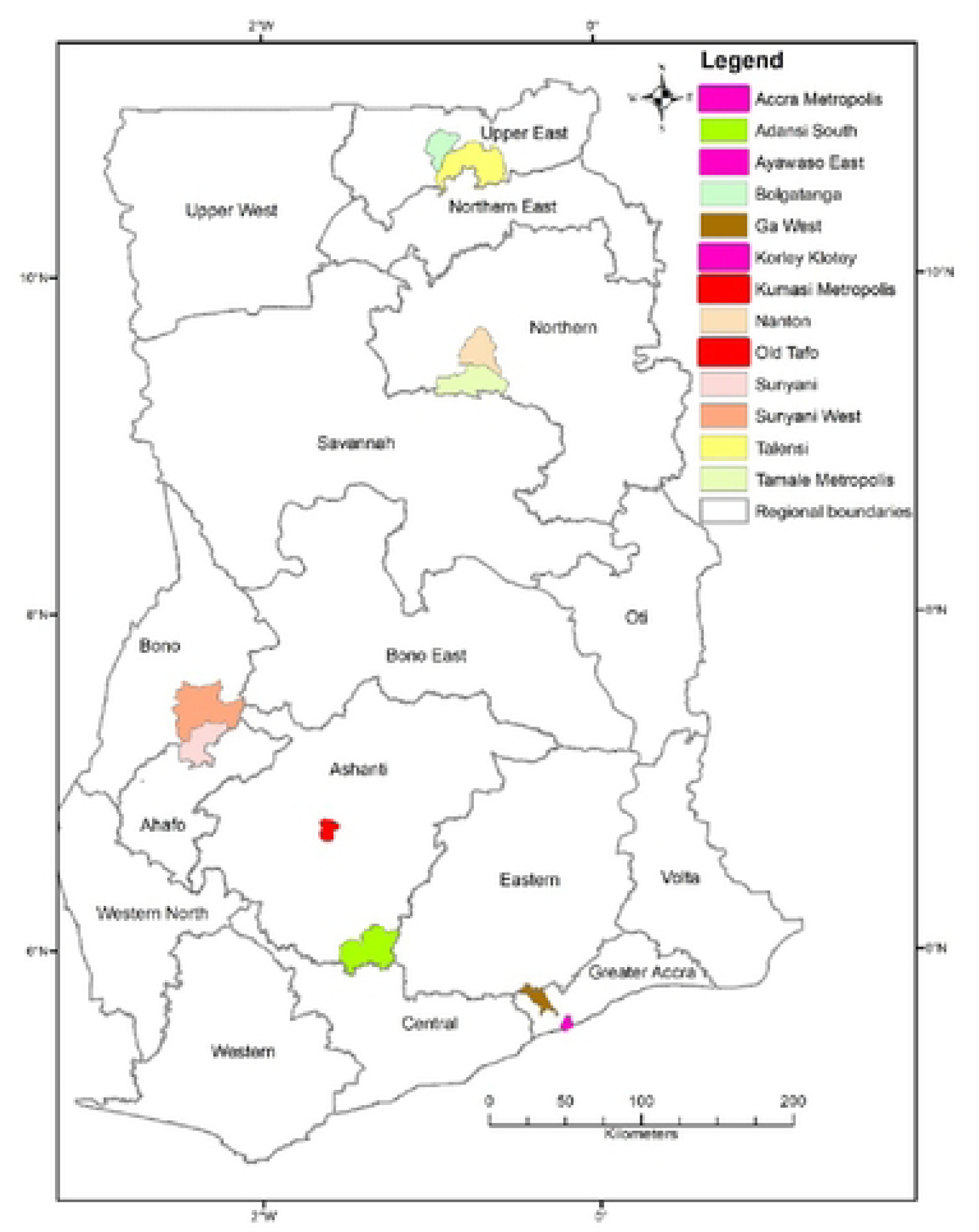
Map of Ghana showing the study locations

In terms of health service delivery, the country operates a five-tiered service system, including: National, Regional, District, Sub-district, and the Community (Ministry of Health, 2011; WHO, 2017). The regional, district and community levels, health facilities are established to provide primary and secondary health care services (WHO, 2017). In 1999, Ghana adopted the Community-based Health Planning and Services (CHPS) strategy as a primary health care system (43–46). In the CHPS strategy, Community Health Officers (CHOs) are assigned to a demarcated CHPS zone(s) as a measure to ensure the close-to-client model of community health services in those demarcated zones (47,48).

### Study Population

The study population included health policy officials, healthcare managers, healthcare service providers, persons with mental health conditions and primary caregivers of people with mental health conditions, civil society organizations, self-advocates, and mental health champions.

### Sample Size Determination

The sample was calculated using Epi Info^TM^ programme https://www.cdc.gov/epiinfo/index.html. The parameters considered were: (i) the size of the population, (ii) prevalence level, (iii) confidence level of 95% and (iv) 5% accepted deviation from expected prevalence and (v) a non-respondent’s rate of 5%. The combine population of the target districts was 956,597 (49). Based on the WHO prevalence rate of 13% (50–53), an estimated 3,095,520 people are living with mental neurological and substance use disorders. The final sample 1010 realised was proportionally split among the target districts to reflect the differences in the populations of those locations.

For the qualitative arm of this study, Key Informant Interviews (KIIs, N=29) of 29 (9 female) respondents, and Focus Group Discussion (FGDs, N=12) involving 75 (40 female) respondents, were conducted (Table 1). After the interview, the interviewee summarized the key issues as a form of participant validation (54). It took between 45-60 minutes to complete an interview session. Both KII and FGDs ended at the point of saturation (55).

**Table 1:**
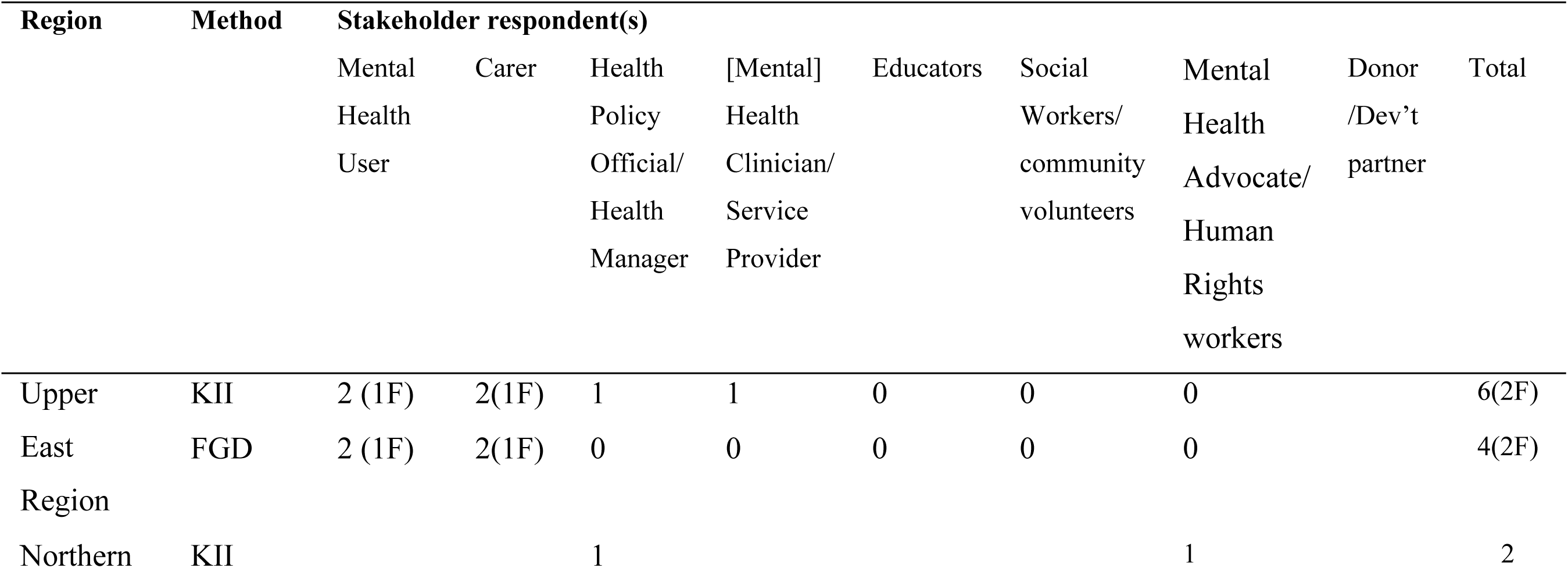
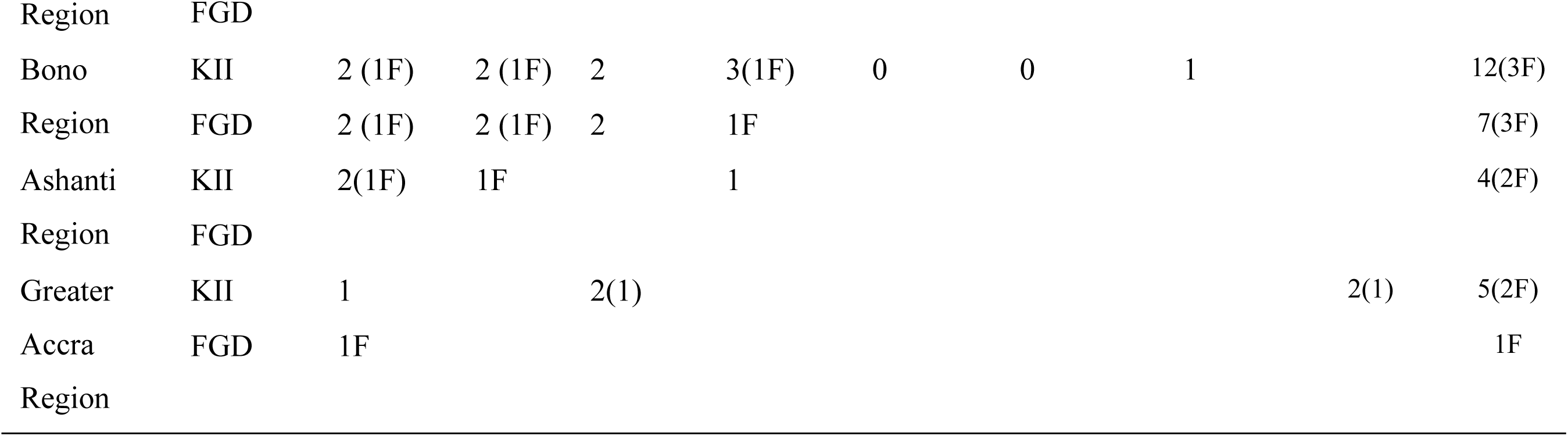
Respondents of qualitative data collection.

### Sampling of Participants

Convenience sampling was used to select stakeholders who have a critical role to play in mental health and were available for data collection (Table 1). The selected individuals were based on initial desk review of available policies and laws on mental health in Ghana and literature in other countries.

### Data Collection Tool

A questionnaire, which was self-administered or interviewer-administered, was used to collect the quantitative data. The governance and leadership topic(s) that were explored covered the level of implementation of legislations and policies on the development and integration of mental health into Primary Health Care, presence of plans, mental health units and focal persons at the PHC, and involvement of other health care staff in mental health services, private sector involvement, and monitoring and evaluation of mental health care services and integration at the PHC. The qualitative data covered the same topics as the questionnaire which allowed for in-depth exploration of the questions.

### Data Collection Procedure

Research assistants were recruited and trained to serve as interviewers to assist the majority of respondents who could not self-complete the questionnaire. Informed consent was obtained for all respondents. Administration of the questionnaires was between 18^th^ August 2020 to 15^th^ December 2020.

The investigators conducted all the KIIs and moderated the FGDs. Triangulation of information collected from the data collected from the different data collection methods was done during analysis to gain richer understanding of complex processes. The qualitative method adopted a critical descriptive ethnography approach. Saturation was achieved when no new and additional information could be obtained. The information was collected over six months between 17^th^ August 2020 and 29^th^ December 2020.

### Analysis

StataSE 16 (64-bit) was used to analyse the quantitative data. Descriptive statistical methods were employed to summarise the data into frequencies and percentage across various leadership and governance domains. The KIIs and FGDs were taped recorded, transcribed verbatim and coded using NVivo 12. The key indicators used during the analysis were: (i) the Mental Health Law and level of awareness at the community level; (ii) mental health policy and level of awareness at the community level; (iii) presence of a mental health plan at the district/ sub-district/ CHPS level and level of implementation; (iv) a mental health unit located at sub-district/ CHPS level; (v) coordinator or focal person(s) for mental health; (vi) involvement of healthcare staff in mental health service development; (vii) private sector participation in mental health service delivery; and (viii) monitoring and evaluation of mental health services.

## Results

A total of 830 study participants completed the survey from a convenience sample of 1010 (response rate of 82%), a majority of them female (56.6%).

By the categories of the survey respondents, a majority were health policy officials and implementers and healthcare service providers, 107 (41.5%), as well as mental health/ human rights advocates, development partners and donors, 54 (48.6%), had high level of awareness of mental health legislation. Awareness of mental health legislation among mental health service users and caregivers was partial with more than half of respondents, (270 [58.6%]), indicating so (Table 2a).

**Table 2a:**
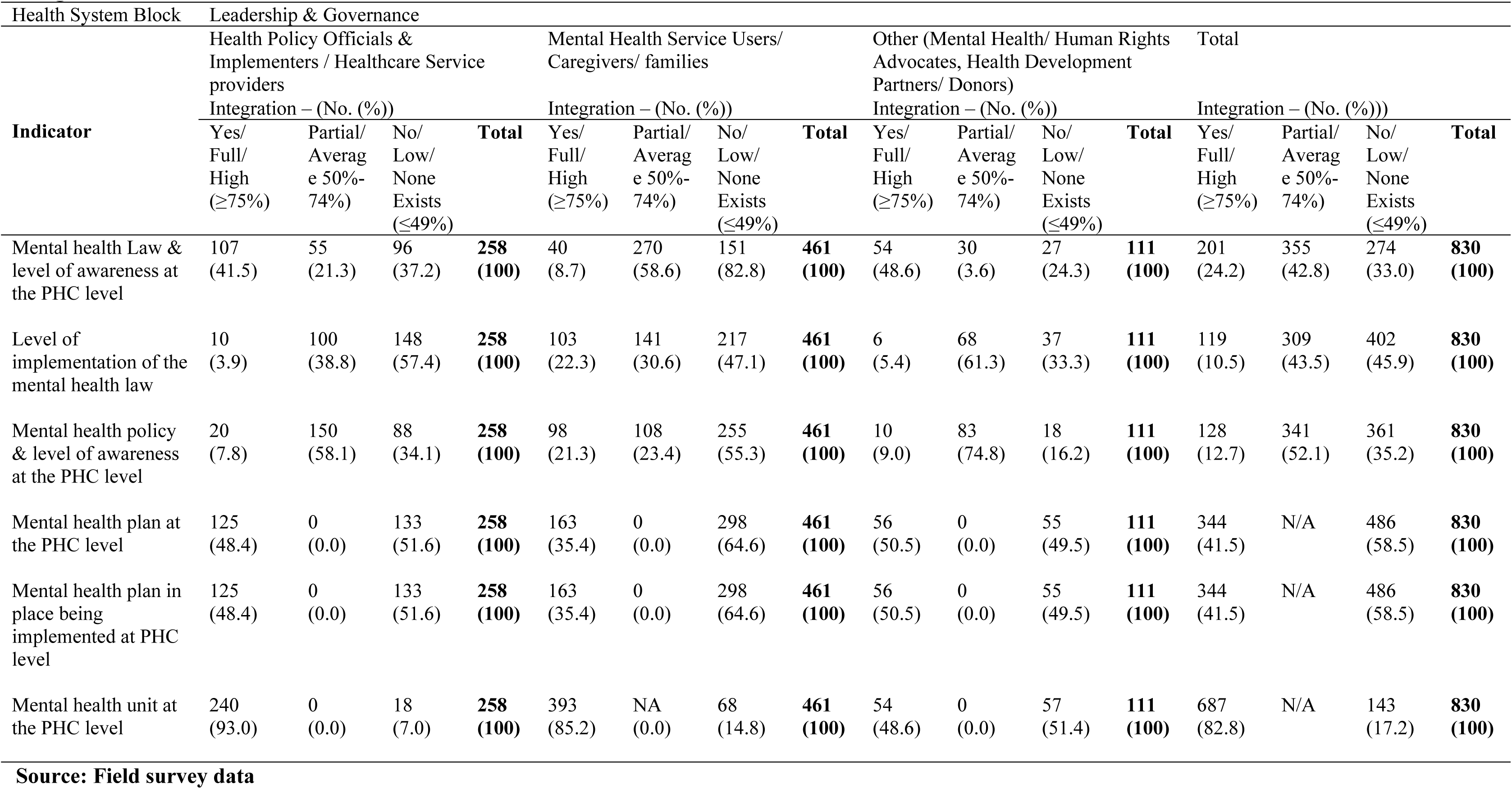
Study participants’ views on mental health leadership and governance at the PHC level and perception of the level of integration in Ghana.

In both the KIIs and FGDs, it emerged that some stakeholders were unaware of the details of the Mental Health Law and level of implementation in the country. The following buttress these points:

> I was part of workshop where they told us about a new mental health law, but I really do not know the details of it or how it can help us. **KII, mental health service user**
>
> We were informed about the mental health law in a meeting that we were called to attend at Dapore-Tindongo [a section of Bolgatanga]. At that meeting we were made to understand that government has a responsibility to look after groups like ours in Bolga[tanga] here. That every person living with mental illness wherever located should be assisted by the government. The law requires of government as such. That is how the shared the information with us at the meeting that we attended. **FGD of mixed group of mental health service users and primary caregivers.**

402 (45.9%) respondents were of the view that the level of implementation of Ghana’s mental health law at the PHC level was low with health policy officials being the largest proportion of respondents (148 (57.4%]) By geographical location of the study respondents, the level of implementation of the mental health law was low among respondents of northern Ghana, 118 (48.0%), and partial for respondents in mid Ghana, by 118 (43.2%), and southern Ghana, 160 (51.4%). Overall, the level of implementation of the mental health law was partial, 380 (45.8%), with 166 (20.0%) of the respondents indicating a high level of implementation (Table 2a).

Results of the qualitative data indicate the level of implementation to be between partial and low:

> “Mental Health Act promoting community-based mental health services – It does promote. Very good. On paper it does promote. In practice, we need to get the structures called for by the Law and that is where I am saying that we need to get the district coordinators in place, which we haven’t got in place yet. So, it leads to a certain gap. We have the regional coordinators they are there, and they are functioning very well, but the district coordinators need to be put in place”. **KII, Health Policy Official**
>
> “There is a mental health law and the NGOs working in mental health and the MHA have done a lot to publicise it, but the law is limited in its compliance with the UNCRPD provisions, particularly the aspect of involuntary treatment. It is at variance with the [UN] CRPD and not completely upholding human rights of people with mental health and psychosocial disabilities”. **KII, mental health advocate.**

More than half, (133 [51.6%]), of health policy officials and health service implementers category indicated that there was no mental health plan at the PHC level. Among the category of mental health service users and caregivers, 298 (64.6%) of them indicated there was no mental health plan in place and or being implemented at the PHC level. For mental health/ human rights advocates however, half of them, (56 [50.5%]), answered in the affirmative that a mental health plan was in place at the PHC level and being implemented. With regards to geographical location of the respondents, the respondents from all three zones were of the view that there was no mental health plan at the PHC level or implemented (Table 2b).

**Table 2b:**
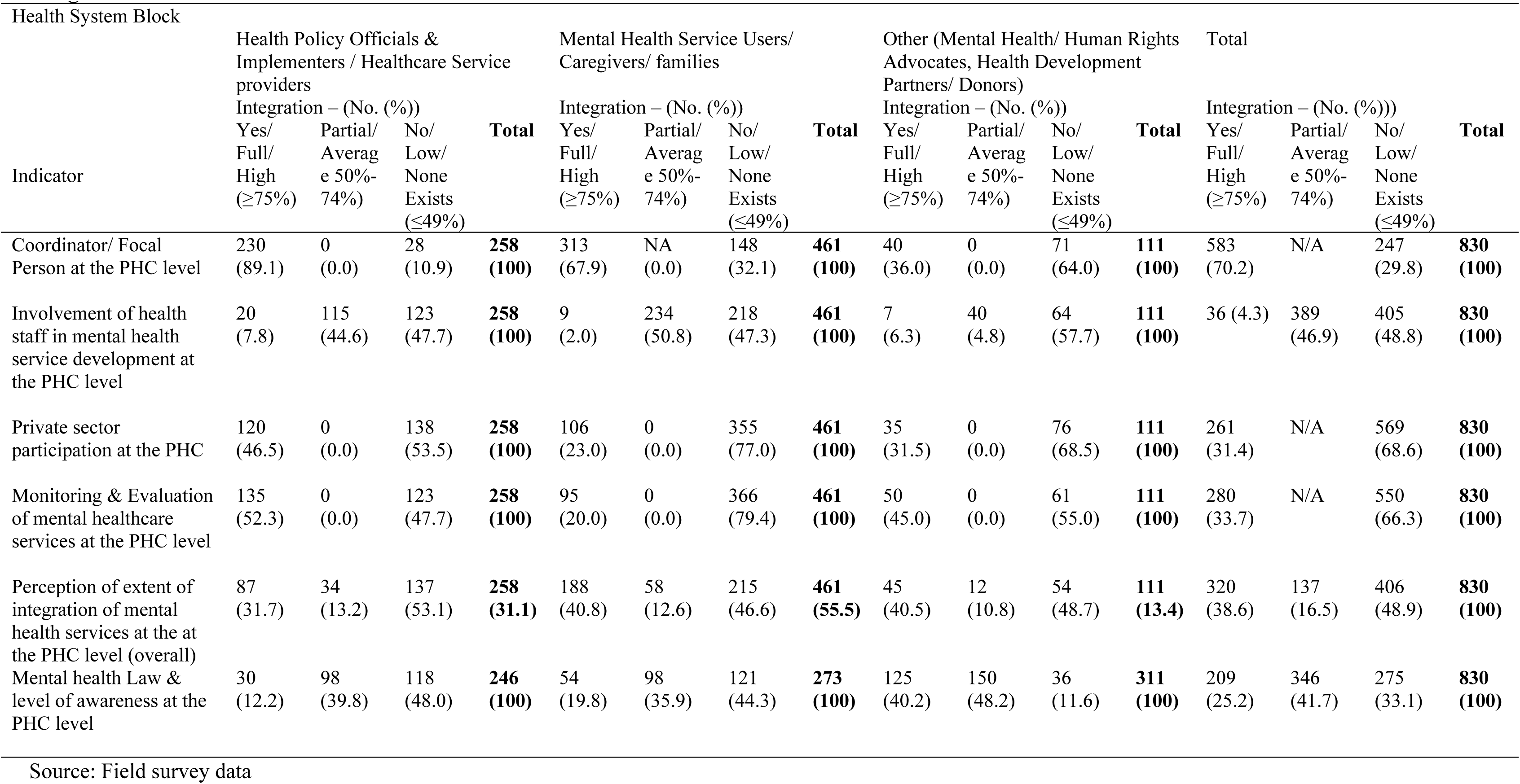
Study participants’ views on mental health leadership and governance at the PHC level and perception of the level of integration in Ghana.

Qualitative data collected, however, suggested that beyond the weekly and month plan of activities that some of the mental health workers draw to work with, there is no annual or strategic plan in place to guide the delivery of mental healthcare services at the district and lower levels of the healthcare system of Ghana.

> *We don’t have plans. Plan of activities depends on finances and since we do not have, we just do the bit we can – some consultations in the hospital for those come and occasional home visits. We only developed plans when NGOs like BasicNeeds* [– Ghana] come to help us or the Mental Health Authority, if they get funding from DFID and channel some to us to work with. **FGD, CPNs/CMHOs, UER.**

The presence of a unit for mental health care at the PHC level is an important indication of a sense of structure and leadership of mental healthcare at that level. Along with interest in knowing if mental health units are present at the PHC, the study also sought to confirm if there was a designated coordinator for mental healthcare at that level. Majority of the respondents answered in the affirmative by 687 (82.8%) for the presence of a mental health unit and 583 (70.3%) for presence of a coordinator or focal person for mental health at the PHC level.

Most respondents were health policy officials and health service implementers, 240 (93.0%) for presence of a mental health unit, and 230 (89.1%) for presence of a mental health coordinator or focal person. Similar responses were provided by mental health service users and caregivers, by 393 (85.2%) for presence of a mental health unit and 313 (67.9%) for presence of a focal person (Table 3a).

**Table 3a:**
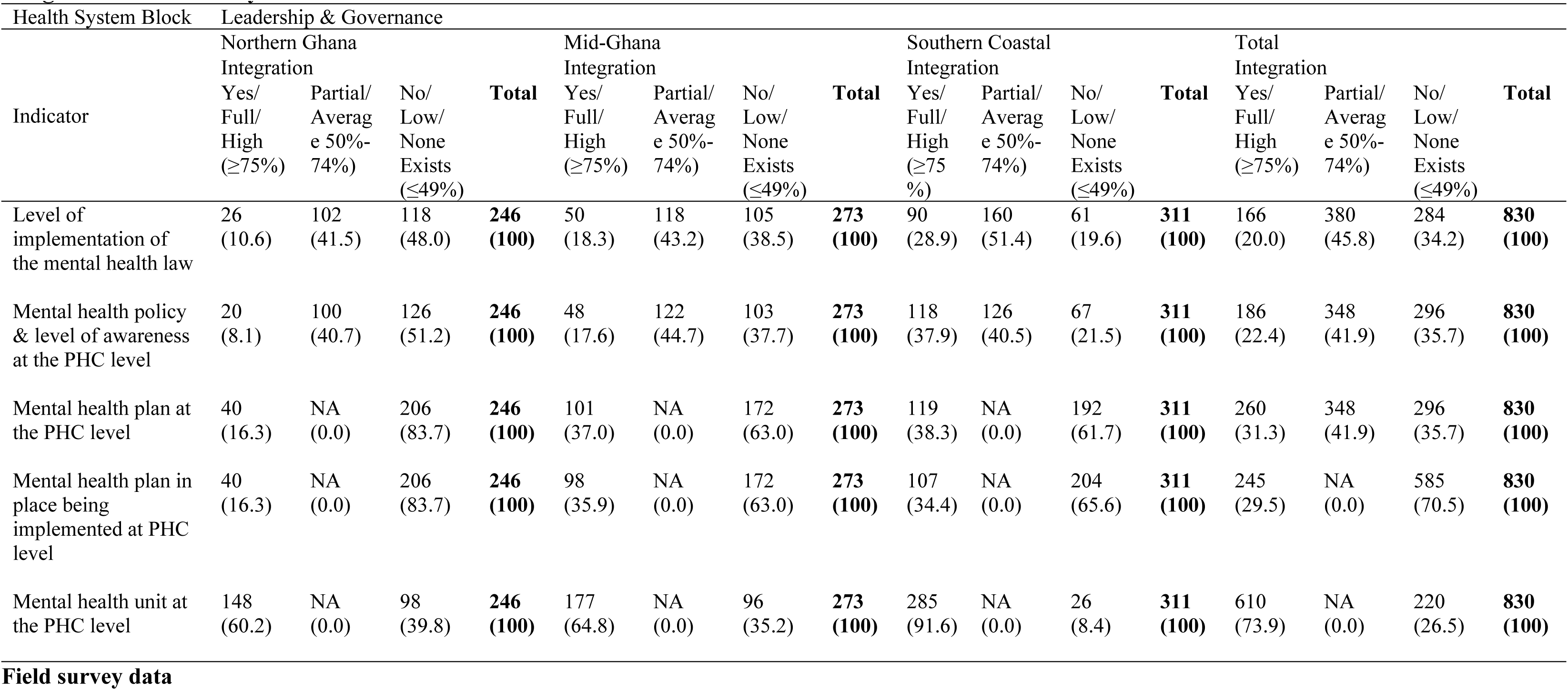
Study participants’ views on mental health leadership and governance at the PHC level and perception of the level of integration in Ghana by location.

However, the difference in percentage of responses indicating the presence of a mental health unit with that for presence of a mental health coordinator/ focal persons could be an indication of units that have no personnel heading or manning them.

Geographically, affirmative responses to the presence of mental health units and mental health coordinators at the PHC level were lower for respondents in northern Ghana, 148 (60.2%), higher in mid Ghana, 179 (65.6%), and highest in, 287 (92.3%), for respondents located in southern Ghana. Responses of the qualitative data collected conveyed mixed views as below:

> At least there is a CMHO in the majority of districts of Ghana but not below. The challenge is the tools and resources for them to work with. I mean funds, logistics like motorbikes, proper offices, and furniture. If these are provided, we can go to the sub-districts and into the hinterlands to provide services. **KII, regional mental health coordinator**
>
> We do not have a unit in our clinic, we have to go to the mental health unit at the regional hospital. **FGD, Service users and caregivers.**
>
> “*You are right that I am a DDNS* [Deputy Director of Nursing Services].*, but I am not one of the Deputy Directors of the Regional Director of Health Services. I am under the DDCC* [Deputy Director of Clinical Care]. So*, I don’t sit at the level to be able to champion mental health. I think the DDCC does his best but mental health will not be his priority. That is what I mean by ‘if your mother is not in the kitchen, you will always be hungry’*” **KII, regional mental health coordinator**.

Absence of representation at the highest level at the regional and national levels of the GHS could be attributed to the low priority placed on mental health. With regards level of involvement of health staff at the PHC level of the healthcare system in mental health service development, a majority of the three categories of survey respondents, 405 (48.8%) were of the view that involvement of healthcare staff was low, followed by 389 (46.9%) who answered that there was a partial level of involvement of health staff in mental health service development and delivery at the PHC level. Just 36 (4.3%) answered that there was a high level of involvement of health staff in mental healthcare development.

Mental health and human rights advocates, development partners and donors were the majority, 64 (57.7%), who indicated that there was a low level of involvement of health staff in mental healthcare service development at the PHC level. Health policy officials and health service implementers, on one hand and mental health service users and caregivers, on the other, respectively, by 123 (47.7%) and 218 (47.3%) expressed that there was low involvement of other health staff in the development of mental health service development in Ghana.

By geographical location, all three zones gauged level of involvement of other health staff in mental healthcare service delivery and integration as low (low, 332 (40.0%) or partial, 300 (36.1%)), with – northern Ghana being the majority, 111 (45.1%), then mid Ghana, 107 (39.2%) and southern Ghana, 114 (36.7%). Qualitative data corroborate the survey results as the quote below convey:

> Unless you are working in mental health, healthcare workers are not interested in mental health matters. There is fear and social stigma. It is only the CPNs [Community Psychiatric Nurses] and the CMHOs [Community Mental Health Officers] who do everything. A negligible number are really interested and involved. **KII, District Director of Health Services**

The study found that private sector participation in the development and delivery of mental healthcare services at the PHC level was low. Private sector participation referred to private clinics and hospitals providing mental healthcare services. Overall, 569 (48.8%) viewed private sector participation in mental, 76 (68.5%), category of mental health and human right advocates, development partners and donors. Health policy officials and health service implementers (138 [53.5%]) held the same view.

For respondents who answered in the affirmative, their reference was the presence of traditional and faith-based healers who are highly patronised by people with mental healthcare needs.

The results of the data analysed by geographical location of the respondents had similar results as the respondent category (Table 3b).

**Table 3b:**
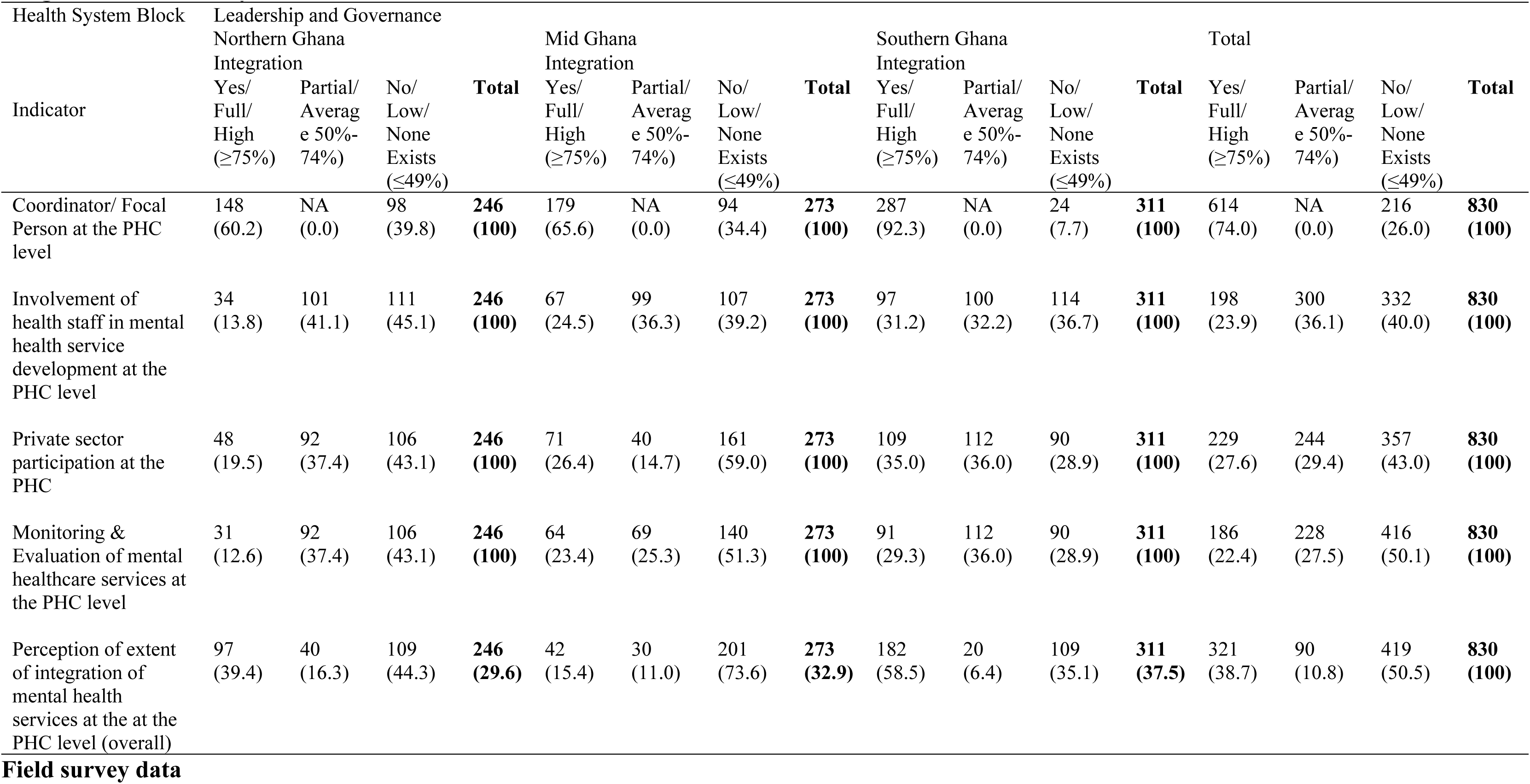
Study participants’ views on mental health leadership and governance at the PHC level and perception of the level of integration in Ghana by location.

Qualitative data analysed corroborate results of the survey, conveyed in the quotes below:

> The services provided by the traditional healers makes me say that there is private sector participation in mental healthcare service delivery in Ghana. There are also a number of the psychiatrists and churches with rehabilitation centres that I have come to be aware of. **KII, Mental health policy official**
>
> For us here, it is only the psychiatry unit that is at the regional hospital. We have many private hospitals and maternity homes, but none offer mental healthcare services. It is only at the government hospitals that mental healthcare services are provided. **FGD, Mental health service users**

Monitoring and evaluation form an important aspect of healthcare stewardship. In this study, Monitoring and Evaluation of mental healthcare services at the PHC level was was assessed to help appreciate leadership and governance of mental healthcare at the community level. Majority of all the three categories of survey respondents combined, 550 (66.3%), were of the view that monitoring and evaluation of mental health care at the PHC level was low. The highest percentage of survey respondents who viewed mental healthcare monitoring and evaluation at the PHC level to be low was the category of mental health service users and caregivers, 366 (79.4%), followed by mental health / human rights advocates, development partners and donors, 61 (55.0%) and finally, health policy officials and health care service providers/ implementers, 123 (47.7%).

By the geographical location of the survey respondents, overall, more than half of them, 416 (50.1%) held the view that monitoring and evaluation at the PHC level was not in place. Only respondents located in southern Ghana, 106 (43.1%), who viewed monitoring and evaluation at the PHC level to be partial. Qualitative data, as per the quote below, lends credence to the results of the survey:

> It was once that staff of the Mental Health Authority at the headquarters and the directors of the Institutional Care Division (ICD) of the Ghana Health Service undertook supportive supervision but that was just to the regional hospital. Also, once a while the Regional Mental Health Coordinator comes around but that is far and in-between. **FGD, CPNs/CHMOs**
>
> Mental health is not considered in matters at this level. supportive supervisions take place, but mental health is not included. We are just sitting and watching. They may include you, but it is not because of mental health but that you an available staff. **KII, CPN**

An overall analysis of the selected leadership and governance indicators which defines stewardship indicate that mental health leadership and governance at the PHC level is low, by 45.2% (375), among the three categories of respondents, and 42.8% (355) by the geographical location of the respondents.

> Leadership is evident when the district director is interested in what is happening with the mental health aspects of his district. Without this level of involvement, we should forget discussing leadership and governance at the district and lower levels. **KII, Mental Health Advocate**

Geographically, respondents in northern Ghana were the highest proportion of respondents, 135 (55.0%) indicating leadership and governance for mental healthcare at the PHC level was low, followed by respondents of mid Ghana, 127 (46.7%) and then respondents of southern Ghana, 92 (29.7%).

> Mental health remains an afterthought and is excluded so leadership and governance is remote, not there. It needs to be consciously crafted and health directors accountable. **KII, National Health Policy**
>
> It is sometimes confusing. You do not know where you belong, who you should be reporting to. So, to be save you report to the [District] Director and then copy the regional mental health coordinator. There hardly any coordination, monitoring or supervision. **FGD, CPNs and CMHOs**

Despite these, there remained significant proportion of respondents who viewed the level of stewardship to be in place by 256 (30.8%) and partial by 199 (24.0%).

> We cannot say there is no leadership and governance of mental health in place at the Primary Care level. Increasingly, directors are taking an interest and through that the situation will improve. **KII, Regional Director of Health Services**

## Discussions

This study was conducted to assess the leadership and governance structures put in place to ensure oversight, coordination, and general stewardship of mental healthcare at the Primary Health Care (PHC) in Ghana. Overall, leadership and governance of mental healthcare at the district and lower levels of the healthcare system of Ghana was average. Just about half of the study respondents were aware of the mental health law and policy and similarly of the view that the level of implementation was partial. Although mental health units and coordinators or focal points for mental health were present in the facilities, their level of involvement in the planning and implementation of healthcare services was minimal, creating mental health silos.

Level of awareness of mental health legislation was affected by the broader social determinants of mental health which was demonstrated in the socio-demographic characteristics of the respondents, particularly geographical locations. The study found that respondents from the northern zone had the lowest level of awareness of mental health law and policy while those from the southern-coastal had the highest level. Challenges with realising the full implementation of the mental health Law (and policy) have been traced to key leadership and governance militating against the development of mental healthcare services in Ghana (56,57). The findings from this current study are consistent with these studies and found that leadership and governance of mental health care at the community level in Ghana remains low.

In comparison to respondents with similar characteristics who were not members of Self-Help peer support Groups (SHGs) that non-governmental organisations such as Basic Needs Ghana support, mental health service users, primary caregivers of people with mental health conditions, and mental health advocates showed a better understanding of current mental-health related policies and legislation. This is a result of the empowerment programs, which are undertaken to increase the participation of those who need mental healthcare services and their carers.

This study found that awareness of mental health laws and policies was only average, indicating that more needs to be done to raise awareness of them, especially among consumers of mental health services, primary caregivers, and other healthcare professionals. Additionally, focal persons find it difficult to understand who is responsible for what under the existing institutional arrangements, which contributes to the apparent inefficiencies. Units are siloed, and district focal persons are both reporting to Mental Health Authority and Ghana Health Service despite the fact that they are at district-level health facilities.

Ghana needs to acknowledge and give priority to the role that mental health plays in the burden of disease, including infectious and non-communicable diseases. This role calls for creative solutions and long-term investments in the core building blocks of the health system framework.

Appropriate policies and legislations are key requirements of the health system framework and Ghana has a mental health law that was passed in 2012 (58) as well as a mental health policy that was launched in 2021 (MoH, 2018). This notwithstanding, Policies and laws alone are insufficient. A national initiative for mental health that is supported by resources, particularly finance, is required. An understanding of the social factors that affect mental health, which take into account regional disparities and associated socioeconomic factors, is necessary for a national mental health programme.

The study revealed that private sector participation was low and when it was identified to exists, it was in reference to the activities of traditional and spiritual healers. This is an indication that the importance of mental health is being recognized gradually but steadily, with the need for decentralized mental healthcare that is incorporated into general healthcare at the PHC level being the best and most obvious solution. This generally average level of awareness This generally average level of awareness suggests that more work needs to be done in order to fully execute the provisions of the mental health law and policy, not only make them known. As an organogram, the primary care level of mental healthcare is generally well established, with the exception of varying service coverage and quality, which are caused by a lack of adequately trained human resources, logistical support, and cross-sectoral linkages. Knowing about the mental health law is one thing, but it is more crucial to know that it is being put into practice. This suggests that increased stakeholder participation in healthcare-related policy debates and legislation is necessary.

In order to achieve the mental health targets that are a part of realizing Universal Health Coverage (UHC) goals and targets, Ghana’s political leadership and healthcare policy authorities must prioritize mental health due to the country’s participation in the Sustainable Development Goals (SDGs). The “epidemiological transition” that many low(-er) and middle-income nations are going through, where mental illness is becoming more prominent and needs to be addressed, is undoubtedly happening in Ghana. The Minister of Health needs to make a strong case to get the President and Cabinet aware of the urgency of addressing the nation’s mental health problems through a thorough implementation of current policies and laws supported by funding to expand access to mental healthcare. A presidential initiative on mental health might help the existing plan to build 111 hospitals in Ghana, including two psychiatric facilities.

Increase awareness of and commitment to implementing existing mental healthcare policy and legislations. Much as this study revealed an appreciably high awareness of existing healthcare policies and legislations, particularly the Mental Health Law, a significant proportion (over 30%) of the population is not aware of those legislations and policies. Coupled with the high levels of stigma and poor attitudes and practices towards mental health and people with mental neurological and substance use illnesses, there is the need to sustain and increase awareness of the legislations and policies.

Health policy makers and community healthcare providers do not generally understand or agree with the inclusion of mental health dimensions on each of the existing healthcare-related policies and legislations on healthcare services, relating to human resources, medicines, and medical products, CHPS, traditional healers, and health insurance and financing. Due to this, mental healthcare is not taken into account in these laws and regulations.

Similar to this, Ghana’s current mental health law and policy are not well understood by healthcare professionals and policymakers who do not work directly in the mental health sub-sector. As a result, the level of attention and responsibility that these individuals should have to support and ensure integration of mental health into general healthcare, especially at the community level, is compromised.. Knowledge and awareness play vital roles in the actual implementation of the provisions and requirements of healthcare policies and legislations. The Ghana Health Service and the Ghana Mental Health Authority must lead in getting their staff and agencies to be well informed and apply policies and legislations, as required. The GHS and MHA should collaborate actively with NGOs/CBOs, especially mental health service-user-led and caregiver associations in the dissemination of policies and legislations and their implementation. The stakeholders involved should consider producing abridged easy-to-read versions of the various healthcare and mental healthcare policies and legislations for wider circulation and dissemination. Inservice-Training (INSET) for healthcare workers and civil society organisations will help increase and imbue into the cross-section of stakeholders and boost advocacy for the implementation and eventual integration of mental health into general healthcare at the primary care level.

## Conclusions

The building blocks of the health systems framework, individually and as a collective, constitute a suitable approach to addressing inadequacies in the development and integration of mental healthcare at the lower levels of the healthcare system in Ghana. Leadership and governance are essential to mental health policy and service development for which they should guide the building of community mental health care system across the country. If the Government of Ghana is to achieve UHC then there must be leadership at the highest level beyond the current efforts of NGOs/CSOs and psychiatrists and psychiatrists, psychiatric nurses, clinical psychologists, and other allied healthcare workers. Such governmental level focus needs to be publicly demonstrated by the ministries of health and finance with a financing and investment mechanism for community mental health.

## Data Availability

All data underlying the findings are available

## Acknowledgement

The authors wish to acknowledge Prof. Patricia Akweongo and Dr Benedict Weobong for their support and advice to the lead author (Peter Badimak Yaro) in his doctoral studies.

## Supporting Information

S1 Figure 1: Map of Ghana showing the study locations

## References

1. Abayneh S, Lempp H, Alem A, Alemayehu D, Eshetu T, Lund C, et al. Service user involvement in mental health system strengthening in a rural African setting: qualitative study. BMC Psychiatry. 2017;17.

2. Faregh N, Lencucha R, Ventevogel P, Dubale BW, Kirmayer LJ. Considering culture, context and community in mhGAP implementation and training: Challenges and recommendations from the field. International Journal of Mental Health Systems. 2019;13(1):1–13.

3. Thornicroft G, Chatterji S, Evans-Lacko S, Gruber M, Sampson N, Aguilar-Gaxiola S, et al. Undertreatment of people with major depressive disorder in 21 countries. British Journal of Psychiatry. 2017;210(2):119–24.

4. Dorsey S, Gray CL, Wasonga AI, Amanya C, Weiner BJ, Belden CM, et al. Advancing successful implementation of task-shifted mental health care in low-resource settings (BASIC): Protocol for a stepped wedge cluster randomized trial. BMC Psychiatry. 2020;20(1):1–14.

5. Eaton J, Gureje O, De Silva M, Sheikh TL, Ekpe EE, Abdulaziz M, et al. A structured approach to integrating mental health services into primary care: Development of the Mental Health Scale Up Nigeria intervention (mhSUN). International Journal of Mental Health Systems. 2018;12(1):1–12.

6. Agyepong IA, Sewankambo N, Binagwaho A, Coll-Seck AM, Corrah T, Ezeh A, et al. The path to longer and healthier lives for all Africans by 2030: the Lancet Commission on the future of health in sub-Saharan Africa. The Lancet. 2017;390(10114):2803–59.

7. Gwaikolo WS, Kohrt BA, Cooper JL. Health system preparedness for integration of mental health services in rural Liberia. BMC Health Services Research. 2017;17(1):1–10.

8. World Health Organization. mhGAP Intervention Guide Mental Health Gap Action Programme Version 2.0 for mental, neurological and substance use disorders in non-specialized health settings. World Health Organizationrganization. 2016;2:1–173.

9. Lund C, Tomlinson M, Patel V. Integration of mental health into primary care in low-and middle-income countries: The PRIME mental healthcare plans. British Journal of Psychiatry. 2016;208:s1–3.

10. World Health Organisation (WHO). Everybody’s business: strengthening health systems to improve health outcomes: WHO’s framework for Action. WHO Production Services, Geneva, Switzerland; 2007. 1–56 p.

11. Member States of the African Region of World Health Organization. The Ouagadougou declaration on primary health care and health systems in Africa: achieving better health for Africa in the new millennium. Brazzaville: WHO Production Services, Geneva, Switzerland; 2008. p. 1–3.

12. Fekadu A, Hanlon C, Medhin G, Alem A, Selamu M, Giorgis TW, et al. Development of a scalable mental healthcare plan for a rural district in Ethiopia. British Journal of Psychiatry. 2016;208:s4–12.

13. WHO. Monitoring the Building Blocks of Health Systems: a Handbook of Indicators and their measurement strategies. Geneva: WHO Document Production Services; 2010. 110 p.

14. Gouda HN, Charlson F, Sorsdahl K, Ahmadzada S, Ferrari AJ, Erskine H, et al. Burden of non-communicable diseases in sub-Saharan Africa, 1990–2017: results from the Global Burden of Disease Study 2017. The Lancet Global Health. 2019;7(10):e1375–87.

15. World Health Organisation. The WHO special initiative for Mental Health (2019-2023) – Universal Coverage for Mental Health. Geneva: WHO; 2019. p. 1–4.

16. WHO. WHO Press. 2022. Health system governance.

17. Upadhaya N, Jordans MJD, Pokhrel R, Gurung D, Adhikari RP, Petersen I, et al. Current situations and future directions for mental health system governance in Nepal: findings from a qualitative study. International Journal of Mental Health Systems. 2017;11:37.

18. USAID. What is health system strengthening? 2021.

19. WHO. WHO Press. 2022. Health system governance.

20. Schneider H, Zulu JM, Mathias K, Cloete K, Hurtig AK. The governance of local health systems in the era of Sustainable Development Goals: reflections on collaborative action to address complex health needs in four country contexts Analysis. BMJ Global Health. 2019;4:1645.

21. Hanlon C, Eshetu T, Alemayehu D, Fekadu A, Semrau M, Thornicroft G, et al. Health system governance to support scale up of mental health care in Ethiopia: a qualitative study. International Journal of Mental Health Systems. 2017;11:38.

22. Buckle G. Realities of leadership and governance in health. 2017 p. 3–39.

23. Republic of Ghana. Ghana Mental Health Act 2012. 2012. p. 44.

24. Walker GH, Osei A. Mental health law in Ghana. 2017;14(2):38–9.

25. Ministry of Health of Ghana. Mental health policy 2019-2030 – Ensuring a mentally health population. Accra, Ghana: Ministry of Health; 2018. 1–90 p.

26. Esponda GM, Hartman S, Qureshi O, Sadler EE, Cohen A, Kakuma R, et al. Barriers and facilitators of mental health programmes in primary care in low-income and middle-income countries. Lancet Psychiatry. 2019;1–15.

27. Hanlon C, Luitel NP, Kathree T, Murhar V, Shrivasta S, Medhin G, et al. Challenges and opportunities for implementing integrated mental health care: a district level situation analysis from five low– and middle-income countries. PLOS One. 2014;9(2):1–12.

28. Oppong S, Kretchy IA, Imbeah EP, Afrane BA. Managing mental illness in Ghana: The state of commonly prescribed psychotropic medicines. International Journal of Mental Health Systems. 2016;10(1):1–10.

29. The Parliament of the Republic of Ghana. Ghana Mental Health Act, 2012 (ACT 846). 846 Ghana: Assembly Press, Accra; 2012 p. 1–44.

30. Parliament of the Republic of a Ghana. Ghana Health Service and Teaching Hospitals (Amendment) Act, 2019 Bill. Ghana; 2019.

31. Badu E, O’Brien AP, Mitchell R. An integrative review of potential enablers and barriers to accessing mental health services in Ghana. Vol. 16, Health Research Policy and Systems. Health Research Policy and Systems; 2018. p. 1–19.

32. Creswell JW. Research design: Qualitaive, Quantitative, and Mixed Methods Approaches. Third. Intercultural Education. Los Angeles: Sage Publications, Inc. 2455 teller Road, CA: Thousand Oaks, CA; 2013. 1–249 p.

33. Creswell JW. Research design: Qualitaive, Quantitative and Mixed Methods Approaches. 4th ed. Vol. 66, Sage Publications, Inc. Los Angeles; 2014. 37–39 p.

34. Creswell JW. Revisiting mixed methods and advancing scientific practices. In: Hesse-Biber SN, Johnson RB, editors. The Oxford handbook of multi-method and mixed methods research inquiry. Oxford, England: Oxford University Press; 2015. p. 57–71.

35. Creswell JW, Creswell JD. Research design: qualitaive, quantitative, and mixed methods approaches. Fifth. Los Angeles: SAGE; 2018. 1–275 p.

36. Georgia State University Library. Research guides: Mixed Methods. 2022.

37. Palinkas LA, Horwitz SM, Green CA, Wisdom JP, Duan N, Hoagwood K, et al. Purposeful sampling for qualitative data collection and analysis in mixed method implementation research. Adm Policy Ment Health. 2015;42(5):533–44.

38. Palaganas EC, Sanchez MC, Molintas VP, Caricativo RD. Reflexivity in Qualitative Research: A Journey of Learning. Qualitative Report. 2017;22(2):426–38.

39. Ye Z, Yang X, Zeng C, Wang Y, Shen Z, Li X, et al. Resilience, Social Support, and Coping as Mediators between COVID-19-related Stressful Experiences and Acute Stress Disorder among College Students in China. Applied Psychology: Health and Well-Being. 2020;

40. Ministry of Health. Ministry of Health Ghana Independent Review Health Sector Programme of Work 2010 Ghana. Accra, Ghana; 2011.

41. World Health Organization. Primary Health Care Systems (PRIMASYS) Case study from Ghana Abridged Version. World Health Organization. Geneva: WHO; 2017. p. 1–48.

42. World Health Organization. Primary Health Care Systems (PRIMASYS) Case study from Ghana Abridged Version. World Health Organization. Geneva: WHO; 2017. p. 1–48.

43. Nyonator FK, Awoonor-Williams JK, Phillips JF, Jones TC, Miller RA. The Ghana Community-based Health Planning and Services Initiative for scaling up service delivery innovation. Vol. 20, Health Policy and Planning. 2005. p. 25–34.

44. Binka FN, Aikins M, Sackey SO, Aryeetey R, Dzodzomenyo M, Esena R, et al. In-depth review of the Community-Based Health Planning Services (CHPS) Programme. Accra, Ghana; 2009.

45. Kweku M, Amu H, Awolu A, Adjuik M, Ayanore MA, Manu E, et al. Community-based health planning and services plus programme in Ghana: A qualitative study with stakeholders in two Systems Learning Districts on improving the implementation of primary health care. PLoS ONE. 2020;15(1):1–24.

46. Abboah-Offei M, Gyasi Darkwa A, Ayim A, Ansah-Ofei AM, Dovlo D, Awoonor-Williams JK, et al. Adapting the Community-based Health Planning and Services (CHPS) to engage poor urban communities in Ghana: Protocol for a participatory action research study. BMJ Open. 2021;11(7):1–9.

47. Awoonor-Williams JK, Bawah AA, Nyonator FK, Rofina A, Oduro A, Ofosu A, et al. The Ghana essential health interventions program: a plausibility trial of the impact of health systems strengthening on maternal & child survival. BMC Health Services Research. 2013;13(Suppl 2):S3.

48. Abboah-Offei M, Gyasi Darkwa A, Ayim A, Ansah-Ofei AM, Dovlo D, Awoonor-Williams JK, et al. Adapting the Community-based Health Planning and Services (CHPS) to engage poor urban communities in Ghana: Protocol for a participatory action research study. BMJ Open. 2021;11(7):1–9.

49. City Population. Ghana: Administrative Divisions. 2020.

50. WHO. WHO | Mental disorders. World Health Organization; 2018.

51. National Academies of Sciences Engineering and Medicine. Providing Sustainable Mental and Neurological Health Care in Ghana and Kenya: Workshop Summary. Washington, DC: National Academies Press. Washington DC: The National Academies Press; 2016.

52. Ohene S. Mental healthcare in Ghana. In: National Academies of Sciences Engineering and Medicine, editor. Providing Sustainable Mental and Neurological Care in Ghana and Kenya: Workshop Summary. Washington DC: National Academies Press; 2016. p. 33–49.

53. Ritchie H, Roser M. Ritchie_Roser_2018_Mental Health_OurWorldInData. Oxford, England: Oxford Martin School, University of Oxford; 2018.

54. Birt L, Scott S, Cavers D, Campbell C, Walter F. Member Checking: A Tool to Enhance Trustworthiness or Merely a Nod to Validation? Qualitative Health Research. 2016;26(13):1802–11.

55. Bowling A. Research Methods in Health: Investigating Health and Health Service. 4th Ed. England: Open University Press; 2014.

56. Doku VCK, Wusu-Takyi A, Awakame J. Implementing the mental health Act in Ghana: any challenges ahead? Ghana Medical Journal. 2012;46(4):241–50.

57. Adu-Gyamfi S. Mental Health Service in Ghana: A Review of the Case. International Journal of Public Health Science (IJPHS). 2017;6(4):299.

58. Walker GH, Osei A. Mental health law in Ghana. 2017;14(2):38–9.

59. Ministry of Health of Ghana. Mental health policy 2019-2030 – Ensuring a mentally health population. Accra, Ghana: Ministry of Health; 2018. 1–90 p.

